# The predictive and prognostic value of peripheral blood antigen-specific memory B cells in phospholipase A2 receptor-associated membranous nephropathy

**DOI:** 10.1101/2023.08.14.23292885

**Authors:** Richard Zhu, Hong Tang, Lilian Howard, Meryl Waldman, Quansheng Zhu

**Author notes:** Corresponding authors: Quansheng Zhu, ImmunoWork, Monrovia, CA, USA, 91016-6353,; Meryl Waldman, Clinical Research Center, NIDDK/Kidney Disease Section, National Institutes of Health, Bethesda, MD, USA 20892-1455.

## Abstract

**Background:** Phospholipase A2 receptor-associated membranous nephropathy (PLA2R-MN) is an anti-PLA2R antibody (PLA2R-Ab) mediated autoimmune kidney disease. Although antibody titer correlates closely with disease activity, whether it can provide longer-term predictions on disease course and progression is unclear. Rituximab, a B-cell depletion therapy, has become the first-line treatment option for PLA2R-MN; however, the response to Rituximab varies among patients.

**Methods:** We developed a flow cytometry-based test that detects and quantifies PLA2R antigen-specific memory B cells (PLA2R-MBCs) in peripheral blood, the primary source for PLA2R-Ab production upon disease relapse. We applied the test to 159 blood samples collected from 28 patients with PLA2R-MN (at diagnosis, during and after immunosuppressive treatment, immunological remission, and relapse) to evaluate the relationship between circulating PLA2R-MBC levels and disease activity.

**Results:** The level of PLA2R-MBCs in healthy controls (n=56) is less than or equal to 1.5% of the total MBC compartment. High circulating PLA2R-MBC levels were detected in two patients post-Rituximab despite achieving immunologic and proteinuric remission, as well as in two patients with negative serum autoantibody but increasing proteinuria. Elimination of these cells with Rituximab improved clinical outcomes. Moreover, five patients exhibited elevated PLA2R-MBC levels before disease relapse, followed by a rapid decline to baseline when relapse became clinically evident. COVID-19 vaccination or SARS-CoV-2 infection significantly affected the dynamics of circulating PLA2R-MBCs.

**Conclusions:** This study suggests that monitoring PLA2R-MBC levels in patients with PLA2R-MN may help refine and individualize immunosuppressive therapy and predict disease course and progression. The technology and findings may also have broader applications in the clinical management of other autoimmune diseases.

## Introduction

Management of autoimmune disease poses numerous clinical challenges, given the unpredictable nature of disease relapses and progression, uncertainty about the appropriate dosage and duration of immunosuppressive treatment with potential for cumulative toxicity, and the lack of effective technology to monitor immunologic activity before tissue injury. Various biomarkers, including autoantibodies, cytokines, microRNAs, and exosomes, have been proposed as a surrogate to monitor autoimmune disease activity^1,2^, but limitations exist. Specifically, these biomarkers generally reflect the severity of tissue injury rather than predicting the course of the disease.

Primary membranous nephropathy (PMN) is a kidney-specific antibody-mediated autoimmune disease characterized by immune deposits in glomerular subepithelial spaces and the presence of proteinuria^3^. Phospholipase A2 receptor (PLA2R) is the most common podocyte antigen in PMN targeted by anti-PLA2R autoantibodies (PLA2R-Abs) in 70-80% of patients^4^. PLA2R-Ab titers typically correlate with disease activity such that serologic remission (antibody depletion) precedes clinical (proteinuric) remission, and reappearance of the antibody signals disease relapse^5,6^. However, whether PLA2R-Ab titers can be used to make longer-term predictions on disease course and progression is unclear. Furthermore, by the time circulating PLA2R-Abs are detectable, relapse and glomerular injury are in progress.

B-cell depletion therapies such as Rituximab have become the first-line treatment option for many autoimmune diseases, including PMN^5^. Rituximab is an anti-human CD20 monoclonal antibody that binds to the B cell lineage and subsequently depletes them from circulation. B cell counts usually recover 6-9 months post-Rituximab therapy and reach normal levels in 9-12 months^7,8^.

Recent studies suggest that the early recovery of memory B cells (MBCs) or class-switched MBCs following Rituximab therapy might predict autoimmune disease relapse, including in idiopathic nephrotic syndrome^8^, rheumatoid arthritis^9^, systemic lupus erythematosus^10^, and others. In addition, high frequencies of MBCs are often associated with poor clinical response to Rituximab^10,11^. However, MBCs are a heterogeneous pool of antigen-experienced class-switched and unswitched B cells, which do not reflect accurately on the dynamics of disease-associated MBCs. Here we investigated the relationship between peripheral blood PLA2R antigen-specific MBC (PLA2R-MBC) levels and disease activity in patients with PLA2R-associated MN (PLA2R-MN).

## Methods

### Patient samples

Deidentified patient peripheral blood mononuclear cells (PBMC) were collected at the Clinical Research Center, National Institute of Diabetes and Digestive and Kidney Diseases/Kidney Disease Section, according to the Institutional Review Board. Written informed consent was obtained in accordance with the Declaration of Helsinki. PBMCs were isolated using Vacutainer^®^ CPT™ mononuclear cell preparation tube (BD Biosciences) and cryopreserved in fetal bovine serum containing 10% DMSO. The cell samples were stored at -80°C prior to flow cytometry analysis. Samples were analyzed in a blinded fashion with respect to clinical status. Unblinding of clinical data occurred only after the completion of flow cytometry analyses. Deidentified cryopreserved healthy control PBMCs were obtained commercially. The ethics committee of ImmunoWork approved the study.

### Peripheral blood B cell isolation and PLA2R-probe staining

Peripheral blood B cells were isolated from PBMCs by negative selection using Dynabeads™ Untouched™ Human B Cells Kit (Invitrogen). The enriched B cells were stained with the PLA2R-probe (Amebtec-PLA2R, ImmunoWork) in PBS (phosphate-buffered saline, pH 7.4) containing 1% bovine serum albumin for 30 minutes at 4°C, followed by 3 washes with PBS and subsequently visualized under a fluorescence microscope (Olympus) at excitation wavelength 490 nm and emission wavelength 525 nm. In separate experiments, the enriched patient B cells were first incubated with the unconjugated PLA2R-epitope complexes (ImmunoWork) for 30 minutes at 4°C, followed by adding PLA2R-probe directly into the cell suspension and further incubated for 30 minutes at 4°C.

### B cell culture and antibody detection

The enriched B cells were cultured in 96-well U-bottom plates in 200 μl IMDM medium containing 10% fetal bovine serum, 100 U/ml penicillin, 100 μg/ml streptomycin, 2 mM L-glutamine, 50 μM 2-mercaptoethanol (all cell culture reagents were from Invitrogen otherwise indicated), supplemented with insulin-transferrin-sodium selenite (insulin, 5 μg/ml; transferrin, 5 μg/ml; sodium selenite, 5 ng/ml; MilliporeSigma), 0.5 μg/ml R848 (Resiquimod, MilliporeSigma), 50 ng/ml IL-21, 1 ng/ml IL-1β, 0.3 ng/ml TNFα, and 1 μg/ml recombinant soluble CD40 ligand multimer (all cytokines were from Miltenyi Biotec) at 37°C in 5% CO_2_, as described previously with modification^12–14^. After two weeks of culture, the supernatants were collected by centrifugation, and the secreted IgG was quantified using IgG (total) human ELISA kit (Invitrogen) following the manufacturer’s instruction. In separate experiments, the supernatants were analyzed using an ELISA plate pre-coated with the purified PLA2R whole extracellular region^15^. The amount of PLA2R protein coated on the ELISA plate was verified by a rabbit anti-human PLA2R polyclonal antibody (MilliporeSigma) diluted in the culture media.

### Flow cytometry

Cryopreserved PBMCs were thawed at 37°C, transferred to the PBMC thawing buffer (1 vial cell per 9 ml buffer at 37°C) (All buffers and reagents were from ImmunoWork otherwise indicated), and incubated for 10 minutes at room temperature. The cells were then collected by centrifugation at 300 g for 5 minutes (room temperature), resuspended in 2 ml of the PBMC staining buffer (room temperature), and enumerated using a cell counter. 1∼2 x 10^6^ cells were aliquoted, centrifuged, and resuspended in 100 μl of ice-cold PBMC staining buffer. The cell suspensions were incubated with the FcR blocker (BD Biosciences) for 10 minutes at 4°C, followed by adding 5 μl of a cocktail containing the fluorochrome-conjugated PLA2R-probe (Alexa488), mouse anti-human CD19 (Pacific blue) and CD27 (APC) antibodies (both from BioLegend) for 30 mins at 4°C. At the end of incubation, cells were vortexed briefly, washed once with 3 ml ice-cold flow cytometry buffer, and collected by centrifugation at 300 g for 5 minutes (4°C). The cells were then resuspended in 300 μl of the ice-cold flow buffer containing 7-AAD (Invitrogen), incubated for 5 minutes on ice in the dark, and proceeded for the flow analysis (BD FACSLyric). 100k events were collected for each PBMC sample. Samples displaying at least 100 events in the CD19^+^CD27^+^ memory B cell compartment were chosen for data analysis. PLA2R-MBCs were gated as CD19^+^CD27^+^PLA2R-probe^+^ population. Cytometry data were analyzed using FlowJo software.

### Statistical analyses

Means ± SD was calculated using SigmaPlot 10 software. Statistical analysis was performed using SigmaPlot 10 software.

## Results and Discussion

### Detection and quantification of PLA2R antigen-specific memory B cells

Antigen-specific MBCs possess unique B cell receptors (BCRs) that react to an antigen in the same manner as the cognate antibody. We therefore conjugated the PLA2R-epitope^15,16^ with a fluorochrome as a probe (PLA2R-probe), to detect and quantify PLA2R-MBCs in peripheral blood (Fig. 1A). We observed that the fluorochrome-conjugated PLA2R-probe produced distinctive staining on the B cells isolated from a patient with PLA2R-MN (in partial remission), but not on the B cells isolated from a healthy control (Fig. 1B). Importantly, the positive staining could be fully blocked by the unconjugated PLA2R-epitope (Fig. 1B), demonstrating the specificity of the PLA2R-probe in detecting PLA2R-MBCs. Furthermore, although the *in vitro* cultured B cells from both subjects secreted significant amounts of IgG antibodies, PLA2R-Abs were detected only in the culture medium of B cells isolated from the patient (Fig. 1C). These observations suggest that the PLA2R-probe-stained B cells are fully functional. We next quantified the PLA2R-MBC levels in the CD19^+^CD27^+^ MBC compartment of peripheral blood mononuclear cells (PBMCs) collected from the patient and healthy control using flow cytometry and PLA2R-probe. We found a high percentage (17.6%) of PLA2R-MBCs in the patient compared to 0.24% in the healthy control (Fig. 1D and Fig. S1).

**Figure 1.**
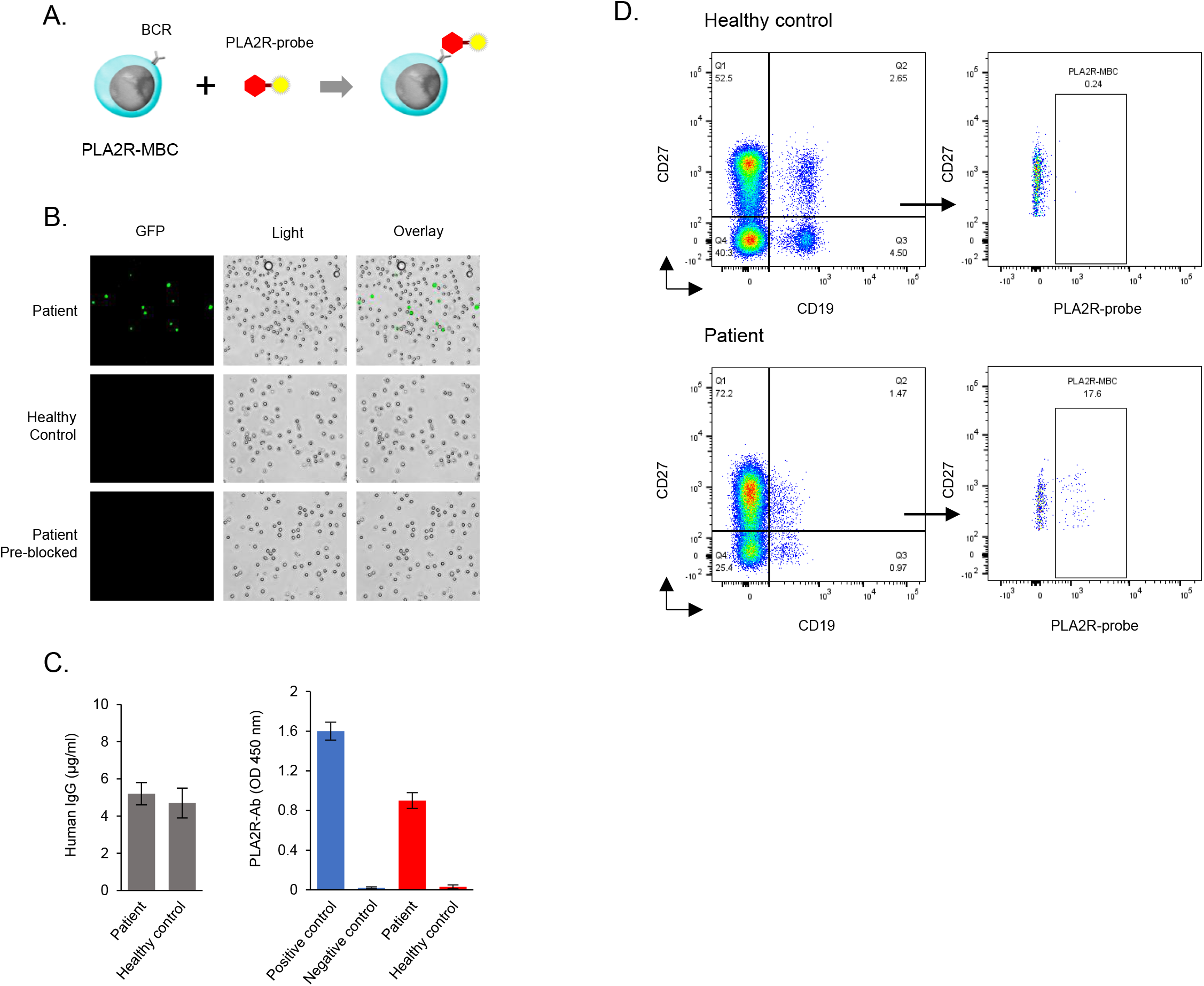
Detection of PLA2R antigen-specific memory B cells using fluorochrome-conjugated PLA2R-probe. A. Cartoon shows the mechanism of PLA2R antigen-specific memory B cell detection. B. Staining of total B cells isolated from a patient with PLA2R-associated membranous nephropathy and a healthy control. The bottom images indicate the pre-blocked patient sample by unconjugated PLA2R-epitope before PLA2R-probe staining. C. Comparison of antibody secretion of in vitro cultured B cells isolated from the patient and healthy control. An equal number of B cells (∼4×10^4^) from both subjects were cultured for 2 weeks before the analysis. A rabbit anti-human PLA2R monoclonal antibody diluted in cell culture media was used as the positive control, and cell culture media alone was used as the negative control for anti-PLA2R antibody ELISA assay. Error bars represent mean ± S.D. (n=3). D. Detection and quantication of PLA2R antigen-specific memory B cells using flow cytometry. The anti-human CD19 and CD27 antibodies were used to indentify the memory B cell compartment. BCR, B cell receptor, GFP, green fluorescent protein channel.

To establish a cutoff value of PLA2R-probe staining in the MBC compartment in the healthy population, we analyzed PBMCs collected from 56 healthy controls using flow cytometry. We observed that 10 samples displayed no staining (0%), 42 samples had staining between 0.05-1.0%, and 4 samples were up to 1.5% (Table 1). As a result, we set the cutoff value of PLA2R-probe staining in healthy controls to less than or equal to 1.5%. We then retrospectively analyzed PBMCs collected serially from 10 MN patients who were PLA2R-Ab negative (based on PLA2R staining on kidney biopsy and serology). As predicted, all patients had low PLA2R-MBC staining values similar to healthy controls regardless of disease stage and treatment (Table 2).

**Table 1.** Characteristics of Healthy controls (n=56).

| Characteristic | n |
| --- | --- |
| Gender |  |
| Male/Female | 34/22 |
| Age (years) |  |
| Mean | 42 |
| Range | 26-72 |
| B cell range (%) | 2.23-13.6 |
| MBC range (%) | 0.56-3.33 |
| PLA2R-MBC range (%) |  |
| 0 | 10 |
| 0.05-1.0 | 42 |
| 1.0-1.5 | 4 |

**Table 2.**
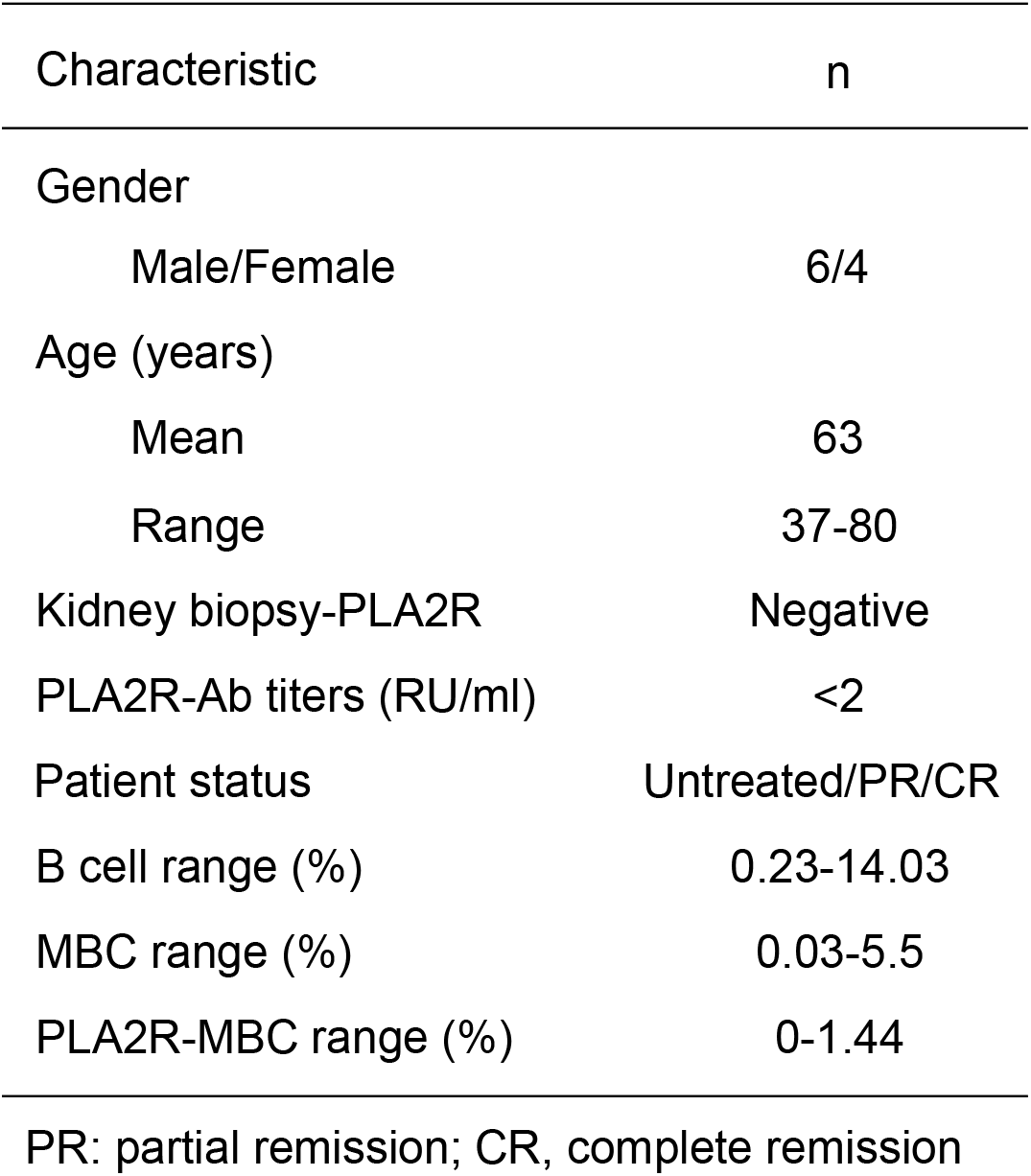
Characteristics of patients with PLA2R-negative MN (n=10).

| Characteristic | n |
| --- | --- |
| Gender |  |
| Male/Female | 6/4 |
| Age (years) |  |
| Mean | 63 |
| Range | 37-80 |
| Kidney biopsy-PLA2R | Negative |
| PLA2R-Ab titers (RU/ml) | <2 |
| Patient status | Untreated/PR/CR |
| B cell range (%) | 0.23-14.03 |
| MBC range (%) | 0.03-5.5 |
| PLA2R-MBC range (%) | 0-1.44 |

PR: partial remission; CR, complete remission

### Association of PLA2R antigen-specific memory B cell levels and disease activity

To evaluate the relationship between PLA2R-MBC levels and disease activity, we longitudinally assessed PLA2R-MBC levels along with PLA2R-Ab titers and proteinuria at different disease stages (at diagnosis, during and after immunosuppressive treatment, at immunological remission, and relapse if relevant) in 19 patients with PLA2R-MN (out of 28 patients analyzed, Table S1). After one cycle of Rituximab, two patients (patients 1 and 2) showed elevated PLA2R-MBC levels (69.5% and 8.77%, respectively) in the repopulated MBC compartment 7 and 12 months after treatment, despite achieving immunologic remission (PLA2R-Ab < 2 RU/ml) and a significant reduction in proteinuria (Fig. 2A, B and Table S2). Retreatment of patient 1 with Rituximab at 7 months (per protocol the patient enrolled in^17^) effectively eliminated the PLA2R-MBCs, and the patient achieved partial remission, maintained for 32 months without evidence of relapse. Considering antigen-specific MBCs are the primary source for cognate antibody production^18^, we reasoned that the second cycle of Rituximab was critical to induce stable disease remission in the patient. In contrast, patient 2, who was not retreated with Rituximab due to seronegative PLA2R-Ab and low proteinuria (partial remission, <0.41 grams/day), had persistently high PLA2R-MBC levels (6.6-10.8%) and relapsed within 2 years. Currently, there is a lack of consensus to guide the optimal number of dosages and courses of Rituximab in patients with MN^5^, to achieve remission and prevent relapse while minimizing exposure to immunosuppression. It is unclear if retreatment decisions should be based on a percentage decrease in PLA2R-Ab titer from baseline and/or, a decrease in proteinuria values, or empirically scheduled after a predetermined period (e.g., 6 months after the first cycle). Our observations suggest that the persistence of a high PLA2R-MBC level after Rituximab therapy, despite negative PLA2R-Ab titers and proteinuric remission, may warrant additional cycles of Rituximab to avoid relapse, whereas those with low PLA2R-MBC levels may not require additional immunosuppression.

**Figure 2.**
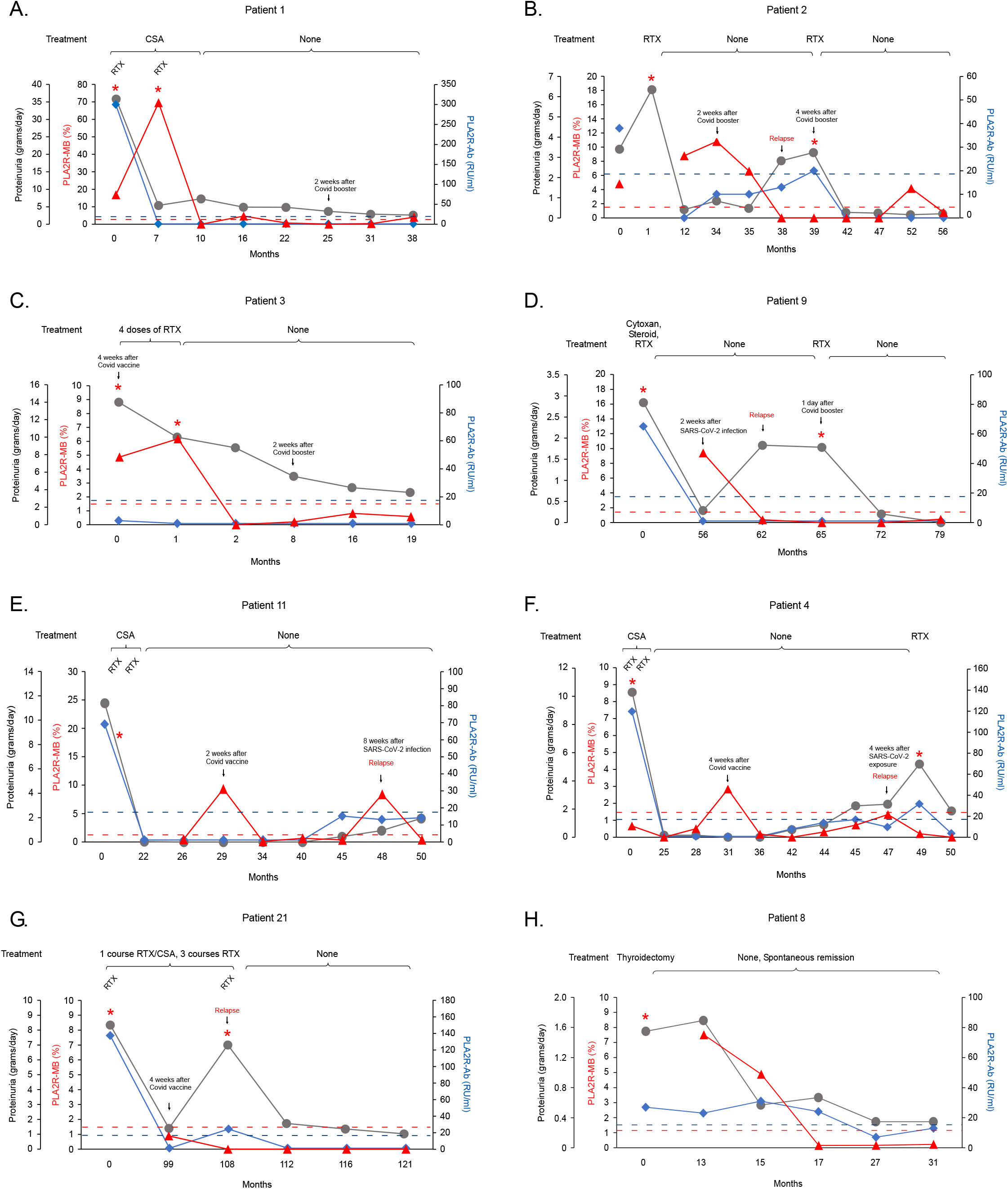
Longitudinal analysis of peripheral blood PLA2R-MBC levels (red triangle) to PLA2R-Ab titers (blue diamond) and proteinuria (grey dot) in patients with PLA2R-associated MN. CSA, Cyclosporine; RTX, Rituximab; *, treatment; dotted lines, cutoff value for PLA2R-Ab (blue, 19 RU/ml) and PLA2R-MBC (red, 1.5%).

Decisions regarding immunosuppressive treatment are also challenging in PMN patients known to be PLA2R associated (based on kidney biopsy or indirect immunofluorescence assay (IFA)), but have serially low or negative PLA2R-Ab titers despite high or increasing proteinuria levels^6^. Lower (or negative) PLA2R-Ab titers often suggest a milder disease course and a greater likelihood of spontaneous remission and thus might not warrant immunosuppression^19^. Patient 3, with a new diagnosis of biopsy-proven PLA2R-MN, highlighted the discordance between persistently low PLA2R-Ab titers (3 RU/ml, IFA+) and sustained high-grade proteinuria (14 grams/day) (Fig. 2C). In this patient, we observed a high circulating PLA2R-MBC level (4.87%) at diagnosis. With one cycle of Rituximab, the PLA2R-MBCs were effectively depleted, along with a substantial decline in proteinuria (3.5 grams/day). The elevated PLA2R-MBC level and high grade of proteinuria in the patient likely indicated an immunologically active disease, which could lead to further disease progression. A similar case was also observed in patient 9, who experienced disease relapse (Fig. 2D). Thus, our findings suggest that a high PLA2R-MBC level might serve as an additional clinical indicator to guide treatment decisions in these patients.

Five of the 8 patients who relapsed during the study period had elevated circulating PLA2R-MBC levels (compared to their established baselines) 3 to 24 months before relapse (Fig. 2B, D, E, F, and Fig. S2S). Four of these patients had a simultaneous elevation in serum PLA2R-Ab titers and proteinuria at relapse (Fig. 2B, E, F, and Fig. S2S), while one remained PLA2R-Ab seronegative despite increasing proteinuria (Fig. 2D). Interestingly, the high PLA2R-MBC level declined rapidly to baseline when relapse became clinically evident based on proteinuria or PLA2R-Ab titer (if detectable). We reasoned that the proliferation of activated PLA2R-MBCs likely caused the elevation of PLA2R-MBC levels before relapse, and the differentiation of PLA2R-MBCs into plasma cells triggered the subsequent rapid decrease in PLA2R-MBC levels. For the 3 patients that relapsed without elevated PLA2R-MBC (Fig. 2G, and Fig. S2P, R), we predict that the extended intervals between sample collection may have missed the time when the PLA2R-MBC levels were high. Our findings suggest that an elevated PLA2R-MBC level in patients after immunosuppressive therapy indicates greater potential for disease relapse, which could serve as an independent biomarker to monitor the earliest sign of PLA2R-MN relapse and guide additional immunosuppression.

Most patients who achieved remission (partial or complete) after Rituximab had consistently low PLA2R-MBC levels concomitant with negative serum PLA2R-Ab. However, transient elevations in PLA2R-MBCs were noted in several patients shortly after COVID-19 vaccination or SARS-CoV-2 infection, and 3 patients experienced disease relapse after the infection (Fig. 2D, E, F). Although causation has yet to be established, glomerular disease relapses associated with SARS-CoV-2 infection and COVID-19 vaccination have been reported^20,21^. We predict that, in the 3 relapsed patients, SARS-CoV-2 infection may have induced the bystander effect^22,23^ that released the PLA2R-MBCs into circulation, which synergized with viral-induced immunologic effects, triggering the disease relapse. We also noted a transient spike in PLA2R-MB levels in 4 patients during the study observation period, yet no proteinuria was detected (Fig. S2J, L, M, S). These observations suggest that the level of circulating PLA2R-MBC in patients is much more dynamic than PLA2R-Ab titers, and may be influenced by viral or other infections, vaccination, or other unknown exposures. Therefore, serial monitoring of PLA2R-MBCs (rather than an absolute value at a single time point) is warranted to determine if the detected PLA2R-MBC level is a transient spike without clinical implications or a persistent elevation with impending disease relapse.

Among 19 PLA2R-MN patients, one coexisted with papillary thyroid cancer (Fig. 2H), presumed to be the cause of MN^24^. Other than thyroidectomy, the patient did not receive immunosuppressive therapy or other medications that decrease proteinuria non-specifically. We observed that, 17 months after surgery, the PLA2R-MBC level declined steeply (from 7.52% to 0%), parallel with a steady decline in proteinuria (1.55 to 0.67 grams/day). Although PLA2R-Ab titer decreased from a peak of 31 to 24 RU/ml, it took additional 10 months before reaching the negative range (<14 RU/ml). The patient had no evidence of relapse 31 months after thyroidectomy. This observation likely reflects the natural course of MN with potential for spontaneous remission in the absence of chronic antigenic stimulation.

Of the 8 newly diagnosed and treatment naïve patients, 4 had a high PLA2R-MBC level (at 16.8%, 4.81%, 4.87%, and 7.52%, respectively), yet the other 4 had a low PLA2R-MBC level similar to healthy controls despite high-grade proteinuria and high titers of PLA2R-Ab (Table S1). Considering that elevated PLA2R-MBC levels occur before proteinuria, as shown in the relapsed patients, we speculate that a high PLA2R-MBC level might have occurred in these patients prior to blood sample collection. On the other hand, a simultaneously elevated PLA2R-MBC level, PLA2R-Ab titer, and proteinuria likely indicate an active and progressive disease course.

Compared to the dynamics of PLA2R-MBC in response to Rituximab therapy, we did not observe a clear association between MBC and B cell levels at 6 months post-Rituximab and disease activity (Table S2). A shorter sample collection interval within 6 months post-Rituximab is likely required to detect such an association^8–10^.

Although human memory B cells are commonly defined through CD19 and CD27 expression^25^, the CD19^+^CD27^+^ lymphocytes also include antibody-secreting plasmablasts^26^ and antigen-activated B cells^27^, which all possess antigen-specific BCRs. As a result, the PLA2R-probe stained CD19^+^CD27^+^ MBCs are likely to be a diverse pool of B lymphocytes expressing PLA2R antigen-specific BCRs. Furthermore, studies have shown that, although Rituximab effectively depletes B cells in circulation, it incompletely depletes or does not affect the switched MBCs in lymph nodes^28,29^. Therefore, we propose that the high peripheral blood PLA2R-MBC level detected in patients post-Rituximab therapy is caused by the migration of the PLA2R-MBCs that were not completely depleted from peripheral lymphoid organs to circulation.

Our study has raised an interesting question on the mechanisms and triggers for autoimmune disease relapse. Although COVID-19 vaccination and certain unknown factors induced PLA2R-MBC elevation in some patients, it was mostly a transient spike without clinical implications. Whereas for SARS-CoV-2 infection-induced PLA2R-MBC elevation, it appeared more likely to lead to the subsequent disease relapse. In addition, a high PLA2R-MBC level could persist for two years before disease relapse, and surgical removal of the thyroid cancer (source of PLA2R autoantigen) led to a spontaneous reduction in PLA2R-MBC level, PLA2R-Ab titer, and proteinuria. It is likely that an autoimmune disease flare depends on the coexistence of at least two essential factors: an elevated antigen-specific MBC level and the availability of autoantigens. We predict that a therapy targeting antigen-specific MBC levels, particularly in patients with a multi-relapsing course, might prevent disease relapse and organ dysfunction.

We acknowledge that our study has a limited sample size, and the observations need to be confirmed in a much larger cohort of patients with extended follow-up. Samples were not collected at regular time intervals, some time points important for data evaluation were missing, and certain factors that may affect B cell homeostasis in patients were not controlled. Nevertheless, the current study assessed for the first time the relationship between antigen-specific memory B cell levels and an autoimmune disease activity longitudinally, which formed a foundation for future more extensive cohort studies.

In summary, the current study uncovered the dynamics of PLA2R-MBCs in patients with PLA2R-MN after the disease onset, during and after immunosuppressive therapies, during immunological remission, and relapse. Our findings suggest that quantifying the PLA2R-MBC levels could provide additional important information to evaluate immunologic disease activity in PLA2R-MN, which may help refine and individualize the dosage and duration of immunosuppressive therapies, predict disease relapse, assess patient status before kidney transplantation, and allow for future treatment strategies specifically targeting antigen-specific memory B cells. The technology and our findings may also have broader applications in other autoimmune diseases in general.

## Data availability

All data pertaining to this article are contained within this article and supplemental data.

## Supporting information

This article contains supporting information.

## Acknowledgments

This study was supported in part by ImmunoWork and the intramural research program of the National Institute of Diabetes and Digestive and Kidney Diseases (to M. W.). We thank all the patients for their willingness to participate in the study.

---

**Supplementary Figure 1.**
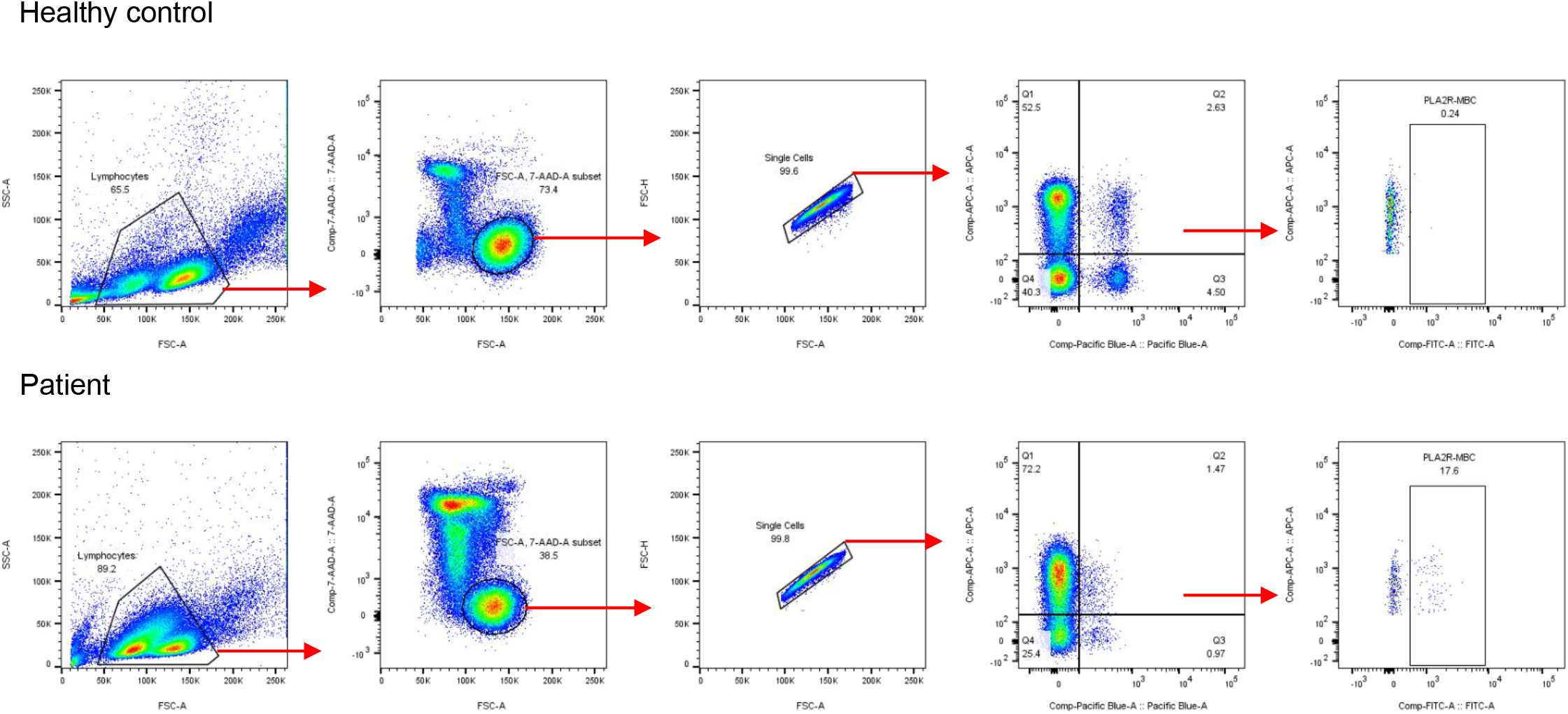
Gating strategy for identifying PLA2R antigen-specific memory B cells. PBMCs (100k events) were first gated by forward (FSC) and side scatter (SSC), extending the normal lymphocyte gate beyond the standard limits into an indivisible group of smaller-sized cell population and a cell population with increased granularity. The live cells were then gated based on 7-AAD staining, followed by comparing the forward scatter area and forward scatter height to exclude the doublets. The single cells were subsequently gated on CD19^+^CD27^+^ cells to obtain the memory B cell population. PLA2R-MBCs were identified as positive stainings in the CD19^+^CD27^+^ B cell population in FITC channel.

**Supplementary Figure 2.**
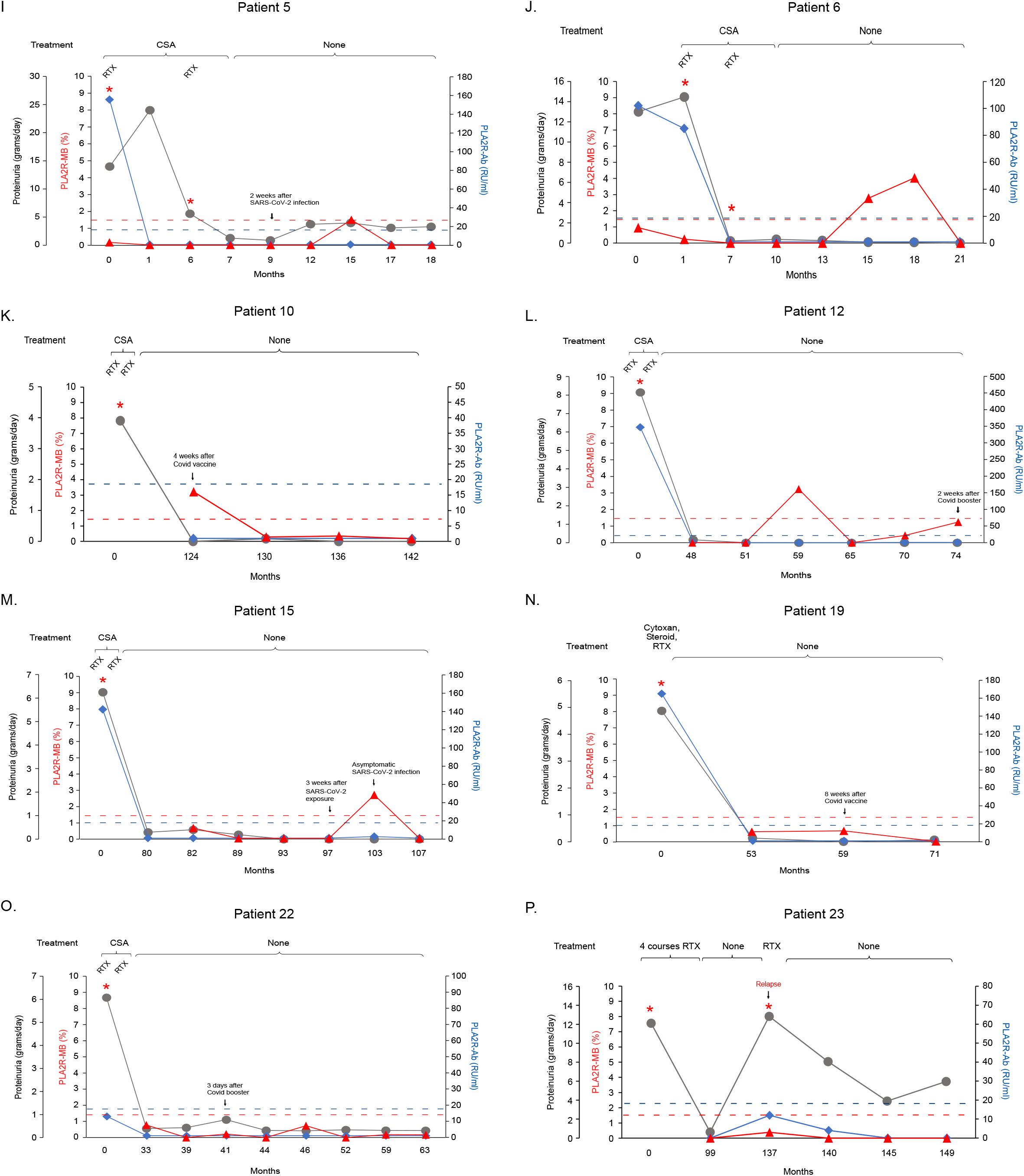
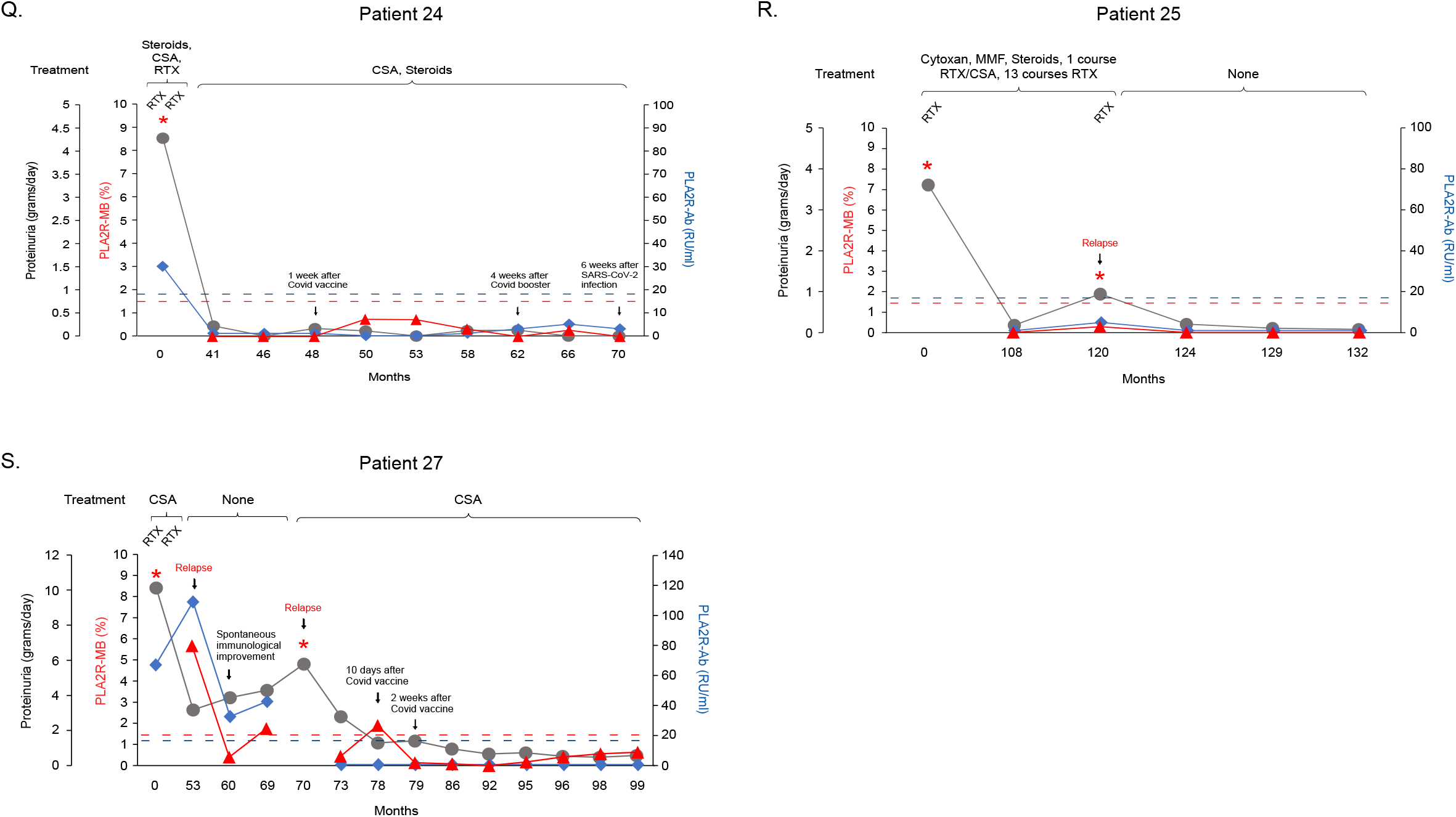

**Supplementary Table 1.** Characteristics of patients with PLA2R-associated MN at first PBMC sample collection.

| Patient ID | Gender | Patient status | Proteinuria (grams/day) | PLA2R-Ab titer (RU/ml) | % B cells in PBMC | % MBC in PBMC | % PLA2R-MBC in total MBC |
| --- | --- | --- | --- | --- | --- | --- | --- |
| 1 | M | New | 35.8 | 300 | 4.03 | 1.64 | 16.8 |
| 2 | M | New | 3.4 | 38 | 5.67 | 2 | 4.81 |
| 3 | F | New | 14 | 3 (IFA+) | 4.79 | 1.84 | 4.87 |
| 4 | M | New | 10.28 | 120 | 6.27 | 1.06 | 0.66 |
| 5 | M | New | 14 | 156 | 3.37 | 2.46 | 0.15 |
| 6 | M | New | 13 | 102 | 8.01 | 3.55 | 0.95 |
| 7* | M | New | 15.6 | 66 | 6.87 | 0.36 | 1.02 |
| 8 | F | New (Thyroidectomy) | 1.69 | 23 | 14.5 | 2.06 | 7.52 |
| 9 | F | CR after treatment | 0.29 | <2 | 6.58 | 0.79 | 9.43 |
| 10 | M | CR after treatment | 0 | <2 | 13.3 | 2.05 | 3.22 |
| 11 | M | CR after treatment | 0 | <2 | 6.07 | 0.14 | 0.44 |
| 12 | F | CR after treatment | 0.16 | <2 | 4.27 | 0.15 | 0 |
| 13* | F | CR after treatment | 0.01 | <2 | 12.5 | 0.54 | 0.25 |
| 14* | M | CR after treatment | 0.28 | <2 | 0.33 | 0.14 | 0 |
| 15 | M | PR after treatment | 0.4 | <2 | 2.01 | 0.32 | 0.64 |
| 16* | M | PR after treatment | 0.57 | <2 | 5.79 | 0.62 | 0 |
| 17* | M | PR on treatment | 0.62 | <2 | 0.15 | 0.08 | - |
| 18* | F | SR | 0.28 | <2 | 4.82 | 0.45 | 8.57 |
| 19 | F | †CR after treatment | 0.14 | <2 | 10.9 | 1.17 | 0.6 |
| 20* | F | †CR after treatment | 0.26 | <2 | 10.2 | 0.53 | 0.48 |
| 21 | M | †PR after treatment | 1.41 | <2 | 2.3 | 0.4 | 0.9 |
| 22 | M | †PR after treatment | 0.39 | <2 | 17.3 | 0.95 | 0.78 |
| 23 | F | †PR after treatment | 0.63 | <2 | 0.06 | 0.04 | - |
| 24 | M | †CR on treatment | 0.22 | <2 | 4.8 | 0.67 | 0 |
| 25 | F | †CR on treatment | 0.19 | <2 | 0.98 | 0.07 | - |
| 26* | M | †PR on treatment | 0.5 | 37 | 2.44 | 1.47 | 17.6 |
| 27 | F | †Relapse | 3.18 | 109 | 6.81 | 1.45 | 5.67 |
| 28* | F | †Relapse | 6.83 | 63 | 5.45 | 0.4 | 1.08 |

PLA2R-Ab titer was measured using the ELISA test from EUROIMMUN. Reference range (RU/mL): <14, negative; 14~19, borderline; >19, positive. Partial remission is defined as protein excretion ≤ 3.5 g/day and ≥ 50% reduction from baseline, complete remission is defined as protein excretion ≤ 0.3 g/day, relapse is defined as the reappearance of proteinuria at least 50% higher in those previously in partial or complete remission and increasing at least on two consecutive evaluations. \*, patient with single sample point; M, male; F, female; New, newly diagnosed patient; CR, complete remission; PR, partial remission; SR, spontaneous remission; †, multi-relapser; IFA+, indirect immunofluorescence test positive; -, MBC counts are too low to calculate.

**Supplementary Table 2.** Detailed serial measurements of B cell, memory B cell, and PLA2R antigen-specific memory B cell populations in patients with PLA2R-associated MN in relationship to clinical parameters, treatments, and additional exposures.

| Patient ID | Time points (months) | Proteinuria (grams/day) | PLA2R-Ab (>19 = positive) | % B cell in lymphocytes | % MBC in lymphocytes | % PLA2R-MBC in total MBC | Treatment and remission status* | SARS-CoV-2 infection or Covid-19 vaccination |
| --- | --- | --- | --- | --- | --- | --- | --- | --- |
| **1** | 0 | 35.85 | 300 | 4.03 | 1.64 | 16.8 | CSA/RTX |  |
|  | 7 | 5.40 | <2 | 1.29 | 0.69 | 69.5 | CSA/RTX |  |
|  | 10 | 7.31 | <2 | 0.044 | 0.028 | - |  |  |
|  | 16 | 4.92 | <2 | 2.21 | 0.26 | 4.62 |  |  |
|  | 22 | 4.89 | <2 | 2.07 | 0.41 | 0.69 |  |  |
|  | 25 | 3.72 | <2 | 2.48 | 0.55 | 0 |  | 2 weeks after Covid-19 booster |
|  | 31 | 2.89 | <2 | 1.98 | 0.79 | 0.3 | PR |  |
|  | 38 | 2.62 | <2 | 2.69 | 0.35 | 3.98 |  |  |
| **2** | 0 | 3.40 | 38 | 5.67 | 2 | 4.81 |  |  |
|  | 1 | 6.34 | NA | NA | NA | NA | RTX |  |
|  | 12 | 0.41 | <2 | 2.77 | 0.27 | 8.77 | PR |  |
|  | 35 | 0.84 | 10 | 6.59 | 0.6 | 10.8 |  | 2 weeks after Covid-19 vaccine |
|  | 36 | 0.48 | 10 | 7.8 | 0.8 | 6.6 |  |  |
|  | 39 | 2.83 | 13 | 7.48 | 1.36 | 0 | Relapse |  |
|  | 40 | 3.23 | 20 | 0.23 | 0.087 | 0 | RTX | 4 weeks after Covid-19 booster |
|  | 43 | 0.28 | <2 | 0.009 | 0.007 | - | CR |  |
|  | 48 | 0.24 | <2 | 0.018 | 0.013 | - |  |  |
|  | 54 | 0.16 | <2 | 2.13 | 0.18 | 4.17 |  |  |
|  | 58 | 0.21 | <2 | 2.25 | 0.22 | 0.76 |  |  |
| **3** | 0 | 14.00 | 3 | 4.79 | 1.84 | 4.87 | RTX | 4 weeks after Covid-19 vaccine |
|  | 1 | 10.00 | <2 | 2.15 | 0.17 | 6.19 | RTX |  |
|  | 2 | 8.79 | <2 | 0.075 | 0.047 | - |  |  |
|  | 8 | 5.53 | <2 | 1.94 | 0.34 | 0.21 |  | 2 weeks after Covid-19 booster |
|  | 16 | 4.24 | <2 | 2.76 | 0.4 | 0.82 |  |  |
|  | 19 | 3.68 | <2 | 3.62 | 0.57 | 0.59 | LR |  |
| **4** | 0 | 10.28 | 120 | 6.27 | 1.06 | 0.66 | CSA/RTX |  |
|  | 25 | 0.14 | <2 | 1.61 | 0.17 | 0 | CR |  |
|  | 28 | 0.14 | <2 | 3.55 | 0.16 | 0.5 |  |  |
|  | 31 | 0 | <2 | 4.01 | 0.3 | 2.84 |  | 4 weeks after Covid-19 vaccine |
|  | 36 | 0 | <2 | 4.67 | 0.45 | 0.18 |  |  |
|  | 42 | 0.54 | 8 | 4.27 | 0.36 | 0 |  |  |
|  | 44 | 0.89 | 14 | 4.04 | 0.5 | 0.31 |  |  |
|  | 45 | 2.23 | 17 | 5.53 | 0.54 | 0.72 |  |  |
|  | 47 | 2.34 | 10 | 3.79 | 0.93 | 1.34 | Relapse | 4 weeks after SARS-CoV-2 exposure |
|  | 49 | 5.19 | 32 | 4.1 | 0.79 | 0.2 | RTX |  |
|  | 50 | 1.86 | 4 | 0.047 | 0.022 | - |  |  |
| **5** | 0 | 14.00 | 156 | 3.37 | 2.46 | 0.15 | CSA/RTX |  |
|  | 1 | 24.00 | <2 | 0.034 | 0.019 | - | CSA |  |
|  | 6 | 5.60 | <2 | 2.01 | 0.088 | - | CSA/RTX |  |
|  | 7 | 1.27 | <2 | 0.004 | 0.004 | - | CSA |  |
|  | 9 | 1.54 | <2 | 0.019 | 0.019 | - | CSA | 2 weeks after SARS-CoV-2 infection |
|  | 12 | 3.74 | <2 | 0.86 | 0.04 | 0 |  |  |
|  | 15 | 3.98 | <2 | 4.2 | 0.14 | 1.49 |  |  |
|  | 17 | 3.11 | <2 | 3.63 | 0.26 | 0 | PR |  |
|  | 18 | 3.30 | <2 | 2.68 | 0.18 | 0 |  |  |
| **6** | 0 | 13.00 | 102 | 8.01 | 3.55 | 0.95 |  |  |
|  | 1 | 14.47 | 85 | 6.34 | 2.67 | 0.24 | CSA/RTX |  |
|  | 7 | 0.19 | <2 | 1.44 | 0.12 | 0 | CSA/RTX |  |
|  | 10 | 0.35 | <2 | 0.049 | 0.006 | - |  |  |
|  | 13 | 0.24 | <2 | 0.04 | 0.015 | - | CR |  |
|  | 15 | 0.08 | <2 | 3.07 | 0.14 | 2.78 |  |  |
|  | 18 | 0.07 | <2 | 4.69 | 0.2 | 4.04 |  |  |
|  | 21 | 0 | <2 | 6.4 | 0.18 | 0 |  |  |
| **8** | 0 | 1.55 | 27 | NA | NA | NA | Thyroidectomy for papillary thyroid cancer |  |
|  | 13 | 1.69 | 23 | 14.5 | 2.06 | 7.52 |  |  |
|  | 15 | 0.57 | 31 | 6.85 | 1.74 | 4.9 | SR |  |
|  | 17 | 0.67 | 24 | 4.45 | 1.39 | 0.16 |  |  |
|  | 27 | 0.35 | 7 | 12 | 1.44 | 0.17 |  |  |
|  | 31 | 0.35 | 13 | 5.07 | 0.83 | 0.23 |  |  |
| **9** | 0 | 2.84 | 65 | NA | NA | NA | Cytosan/ Steroids/RTX |  |
|  | 56 | 0.29 | <2 | 6.58 | 0.79 | 9.43 | CR | 2 weeks after SARS-CoV-2 infection |
|  | 62 | 1.83 | <2 | 3.1 | 0.91 | 0.39 | Relapse |  |
|  | 65 | 1.79 | <2 | 8.12 | 0.89 | 0 | RTX | 1 day after Covid-19 booster |
|  | 72 | 0.20 | <2 | 0.22 | 0.082 | - | CR |  |
|  | 79 | 0 | <2 | 2.07 | 0.48 | 0.47 |  |  |
| **10** | 0 | 0.39 | PLA2R-Ab positive by Western-blot | NA | NA | NA | CSA/RTX |  |
|  | 124 | 0 | <2 | 13.3 | 2.05 | 3.22 | CR | 4 weeks after Covid-19 vaccine |
|  | 130 | 0.09 | <2 | 5.31 | 1.01 | 0.29 |  |  |
|  | 136 | 0 | <2 | 3.95 | 0.58 | 0.35 |  |  |
|  | 142 | 0 | <2 | 5.72 | 1 | 0.18 |  |  |
| **11** | 0 | 11.40 | 69 | NA | NA | NA | CSA/RTX |  |
|  | 22 | 0.04 | <2 | NA | NA | NA | CR |  |
|  | 26 | 0 | <2 | 6.07 | 0.14 | 0.44 |  |  |
|  | 29 | 0 | <2 | 10.8 | 0.36 | 9.3 |  | 2 weeks after Covid-19 vaccine |
|  | 34 | 0 | <2 | 5.98 | 1.44 | 0.067 |  |  |
|  | 40 | 0 | <2 | 6.86 | 0.52 | 0.61 |  |  |
|  | 45 | 0.44 | 15 | 8.88 | 1.14 | 0.33 |  |  |
|  | 48 | 0.94 | 13 | 5.48 | 1.03 | 8.37 | Relapse | 8 weeks after SARS-CoV-2 infection |
|  | 50 | 1.93 | 14 | 4.9 | 1 | 0.37 |  |  |
| **12** | 0 | 8.15 | 347 | NA | NA | NA | CSA/RTX |  |
|  | 48 | 0.16 | <2 | 4.27 | 0.15 | 0 | CR |  |
|  | 51 | 0 | <2 | 8.56 | 0.25 | 0 |  |  |
|  | 59 | 0 | <2 | 5.62 | 0.39 | 3.23 |  |  |
|  | 65 | 0 | <2 | 19.2 | 0.8 | 0 |  |  |
|  | 70 | 0 | <2 | 6.79 | 0.45 | 0.43 |  |  |
|  | 74 | 0 | <2 | 3.15 | 0.28 | 1.24 |  | 2 weeks after Covid-19 booster |
| **15** | 0 | 6.25 | 142 | NA | NA | NA | CSA/RTX |  |
|  | 80 | 0.30 | <2 | NA | NA | NA |  |  |
|  | 82 | 0.40 | <2 | 2.01 | 0.32 | 0.64 |  |  |
|  | 89 | 0.19 | <2 | 2.13 | 0.29 | 0 | CR |  |
|  | 93 | 0 | <2 | 2.79 | 0.35 | 0 |  |  |
|  | 97 | 0 | <2 | 2.18 | 0.31 | 0 |  | 3 weeks after SARS-CoV-2 exposure |
|  | 103 | 0 | 2.71 | 3.2 | 0.57 | 2.71 |  | Asymptomatic SARS-CoV-2 infection |
|  | 107 | 0 | <2 | 3.66 | 0.52 | 0 |  |  |
| 19 | 0 | 4.87 | 165 | NA | NA | NA | Cytoxan/Steroids/RTX |  |
|  | 53 | 0.14 | <2 | 10.9 | 1.17 | 0.6 | CR |  |
|  | 59 | 0 | <2 | 9.06 | 1.25 | 0.65 |  | 8 weeks after Covid-19 vaccine |
|  | 71 | 0.08 | <2 | 6.83 | 0.79 | 0 |  |  |
| 21 | 0 | 8.34 | 137 | NA | NA | NA | CSA/RTX |  |
|  | 99 | 1.41 | <2 | 2.3 | 0.4 | 0.9 | PR | 4 weeks after Covid-19 vaccine |
|  | 108 | 7.01 | 24 | 2.6 | 0.17 | 0 | Relapse, RTX |  |
|  | 112 | 1.73 | <2 | 0.012 | 0.012 | - | PR |  |
|  | 116 | 1.36 | <2 | 0.13 | 0.061 | - |  |  |
|  | 121 | 1.03 | <2 | 2.58 | 0.18 | 0 |  |  |
| 22 | 0 | 6.08 | 13 | NA | NA | NA | RTX/CSA |  |
|  | 33 | 0.39 | <2 | 17.3 | 0.95 | 0.78 | PR |  |
|  | 39 | 0.43 | <2 | 16.8 | 0.99 | 0 |  |  |
|  | 41 | 0.78 | <2 | 21.3 | 0.68 | 0.22 |  | 3 days after Covid-19 vaccine |
|  | 44 | 0.31 | <2 | 9.31 | 0.72 | 0 |  |  |
|  | 46 | 0.30 | <2 | 26.1 | 1.16 | 0.73 |  |  |
|  | 52 | 0.34 | <2 | 9.74 | 0.9 | 0 |  |  |
|  | 59 | 0.31 | <2 | 13.6 | 1.24 | 0.18 |  |  |
|  | 63 | 0.31 | <2 | 11.2 | 1.4 | 0.17 |  |  |
| 23 | 0 | 12.10 | PLA2R-Ab positive by Western-Blot | NA | NA | - | 1 month after RTX |  |
|  | 99 | 0.63 | <2 | 0.062 | 0.035 | - | PR |  |
|  | 137 | 12.83 | 12 | 4.73 | 0.43 | 0.39 | Relapse, RTX |  |
|  | 140 | 8.06 | 4 | 0.002 | 0.002 | - |  |  |
|  | 145 | 3.90 | <2 | 0.07 | 0.037 | - |  |  |
|  | 149 | 5.96 | <2 | 2.62 | 0.087 | - |  |  |
| 24 | 0 | 4.28 | 30 | NA | NA | NA | Steroids/CSA/RTX |  |
|  | 41 | 0.22 | <2 | 4.8 | 0.67 | 0 | Steroids/CSA, CR |  |
|  | 46 | 0 | <2 | 9.74 | 0.74 | 0 | " |  |
|  | 48 | 0.16 | <2 | 4.86 | 0.59 | 0 | " | 1 week after Covid-19 vaccine |
|  | 50 | 0.11 | <2 | 11.1 | 1.22 | 0.74 | " |  |
|  | 53 | 0 | <2 | 4.73 | 0.63 | 0.73 | " |  |
|  | 58 | 0.12 | <2 | 3.67 | 0.62 | 0.32 | " |  |
|  | 62 | 0.13 | 3 | 7.77 | 1.34 | 0 | " | 4 weeks after Covid-19 booster |
|  | 66 | 0 | 5 | 4.23 | 0.77 | 0.27 | " |  |
|  | 70 | 0 | 3 | 4.26 | 0.8 | 0 | " | 6 weeks after SARS-CoV-2 infection |
| **25** | 0 | 3.61 | PLA2R-Ab positive by Western-blot | NA | NA | NA | Cytosan/MMF/ Steroids/CSA/ RTX |  |
|  | 108 | 0.19 | <2 | 0.98 | 0.069 | - | CSA/steroids |  |
|  | 120 | 0.95 | 5 | 8.28 | 0.78 | 0.28 | Relapse, RTX |  |
|  | 124 | 0.21 | <2 | 0.045 | 0.037 | - | CR |  |
|  | 129 | 0.11 | <2 | 0.086 | 0.019 | - |  |  |
|  | 132 | 0.08 | <2 | 1.11 | 0.1 | 0 |  |  |
| **27** | 0 | 10.13 | 67.2 | NA | NA | NA | CSA/RTX |  |
|  | 53 | 3.18 | 109 | 6.81 | 1.45 | 5.67 | Relapse |  |
|  | 60 | 3.87 | 33 | 10.1 | 2.69 | 0.4 | Spontaneous immunological improvement |  |
|  | 69 | 4.30 | 43 | 11.9 | 3.04 | 1.75 |  |  |
|  | 70 | 5.79 | NA | NA | NA | NA | Relapse, CSA |  |
|  | 73 | 2.79 | <2 | 2.91 | 1.53 | 0.44 | " |  |
|  | 78 | 1.30 | <2 | 5.67 | 1.67 | 1.9 | " | 10 days after Covid- 19 vaccine |
|  | 79 | 1.41 | <2 | 9.49 | 3.75 | 0.13 | " | 2 weeks after Covid-19 vaccine |
|  | 86 | 0.95 | <2 | 7.1 | 2.83 | 0.08 | " |  |
|  | 92 | 0.67 | <2 | 7.63 | 2.83 | 0 | " |  |
|  | 95 | 0.73 | <2 | 6.77 | 3.24 | 0.17 | " |  |
|  | 96 | 0.53 | <2 | 7.41 | 2.95 | 0.41 | " |  |
|  | 98 | 0.48 | <2 | 4.81 | 2.12 | 0.56 | " |  |
|  | 99 | 0.57 | <2 | 6.79 | 2.58 | 0.63 | " |  |

PLA2R-Ab titer was measured using the ELISA test from EUROIMMUN. Reference ranges (RU/ml): <14, negative; 14~19, borderline; >19, positive. Partial remission is defined as protein excretion ≤ 3.5 grams/day and ≥ 50% reduction from baseline, complete remission is defined as protein excretion ≤ 0.3 grams/day, relapse is defined as reappearance of proteinuria at least 50% higher in those previously in partial or complete remission and increasing at least on two consecutive evaluations, limited response is defined as >50% reduction in protein excretion from baseline by >3.5 grams/day. \*, remission status is based on proteinuria, blank cells in the column indicate patients are off immunosuppressants with no change in status compared to previous time point; NA, time points are unavailable; -, MBC counts are too low to calculate; RTX, Rituximab; CSA, Cyclosporine; MMF, Mycophenolate mofetil; PR, partial remission; CR, complete remission; SR, spontaneous remission; LR, limited response; ", on immunosuppressive treatment as indicated above.

## References

1 Fenton KA, Pedersen HL. Advanced methods and novel biomarkers in autoimmune diseases -a review of the recent years progress in systemic lupus erythematosus. Front Med (Lausanne) 2023; 10. DOI:10.3389/fmed.2023.1183535.

2 Xu K, Liu Q, Wu K, et al. Extracellular vesicles as potential biomarkers and therapeutic approaches in autoimmune diseases. J Transl Med 2020; 18: 432.

3 Ronco P, Beck L, Debiec H, et al. Membranous nephropathy. Nat Rev Dis Primers 2021; 7: 69.

4 Beck LH, Bonegio RGB, Lambeau G, et al. M-Type phospholipase A2 receptor as target antigen in idiopathic membranous nephropathy. New England Journal of Medicine 2009; 361: 11–21.

5 Gauckler P, Shin J Il, Alberici F, et al. Rituximab in membranous nephropathy. Kidney Int Rep 2021; 6: 881–93.

6 Lerner GB, Virmani S, Henderson JM, Francis JM, Beck LH. A conceptual framework linking immunology, pathology, and clinical features in primary membranous nephropathy. Kidney Int 2021; 100: 289–300.

7 Roll P, Palanichamy A, Kneitz C, Dorner T, Tony H-P. Regeneration of B cell subsets after transient B cell depletion using anti-CD20 antibodies in rheumatoid arthritis. Arthritis Rheum 2006; 54: 2377–86.

8 Colucci M, Carsetti R, Cascioli S, et al. B cell reconstitution after rituximab treatment in idiopathic nephrotic syndrome. Journal of the American Society of Nephrology 2016; 27: 1811–22.

9 Leandro MJ, Cambridge G, Ehrenstein MR, Edwards JCW. Reconstitution of peripheral blood B cells after depletion with rituximab in patients with rheumatoid arthritis. Arthritis Rheum 2006; 54: 613–20.

10 Vital EM, Dass S, Buch MH, et al. B cell biomarkers of rituximab responses in systemic lupus erythematosus. Arthritis Rheum 2011; 63: 3038–47.

11 Reddy V, Klein C, Isenberg DA, et al. Obinutuzumab induces superior B-cell cytotoxicity to rituximab in rheumatoid arthritis and systemic lupus erythematosus patient samples. Rheumatology 2017; 56: 1227–37.

12 Bezstarosti S, Kramer CSM, Franke-van Dijk MEI, et al. HLA-DQ-Specific recombinant human monoclonal antibodies allow for in-depth analysis of HLA-DQ epitopes. Front Immunol 2021; 12: 761893.

13 Lighaam LC, Vermeulen E, den Bleker T, et al. Phenotypic differences between IgG4+ and IgG1+ B cells point to distinct regulation of the IgG4 response. Journal of Allergy and Clinical Immunology 2014; 133: 267–270.e6.

14 Naito M, Hainz U, Burkhardt UE, et al. CD40L-Tri, a novel formulation of recombinant human CD40L that effectively activates B cells. Cancer Immunology, Immunotherapy 2013; 62: 347–57.

15 Tang H, Zhu R, Waldman M, Zhu Q. Structural determinants of the dominant conformational epitopes of phospholipase A2 receptor in primary membranous nephropathy. Journal of Biological Chemistry 2022; 298: 101605.

16 Kao L, Lam V, Waldman M, Glassock RJ, Zhu Q. Identification of the immunodominant epitope region in phospholipase A2 receptor-mediating autoantibody binding in idiopathic membranous nephropathy. Journal of the American Society of Nephrology 2015; 26: 291–301.

17 Waldman M, Beck LH, Braun M, Wilkins K, Balow JE, Austin HA. Membranous nephropathy: pilot study of a novel regimen combining cyclosporine and Rituximab. Kidney Int Rep 2016; 1: 73–84.

18 Phad GE, Pinto D, Foglierini M, et al. Clonal structure, stability and dynamics of human memory B cells and circulating plasmablasts. Nat Immunol 2022; 23: 1076–85.

19 De Vriese AS, Glassock RJ, Nath KA, Sethi S, Fervenza FC. A proposal for a serology-based approach to membranous nephropathy. Journal of the American Society of Nephrology 2017; 28: 421–30.

20 Klomjit N, Alexander MP, Fervenza FC, et al. COVID-19 vaccination and glomerulonephritis. Kidney Int Rep 2021; 6: 2969–78.

21 May RM, Cassol C, Hannoudi A, et al. A multi-center retrospective cohort study defines the spectrum of kidney pathology in Coronavirus 2019 Disease (COVID-19). Kidney Int 2021; 100: 1303–15.

22 Kardava L, Rachmaninoff N, Lau WW, et al. Early human B cell signatures of the primary antibody response to mRNA vaccination. Proceedings of the National Academy of Sciences 2022; 119: e2204607119.

23 Horns F, Dekker CL, Quake SR. Memory B cell activation, broad anti-influenza antibodies, and bystander activation revealed by single-cell transcriptomics. Cell Rep 2020; 30: 905–913.e6.

24 Zirino F, Gembillo G, Lamanna F, et al. SAT-367 A case of thyroid papillary carcinoma associated with membranous anti-PLA2R positive glomerulonephritis. Kidney Int Rep 2020; 5: S154–5.

25 Sanz I, Wei C, Jenks SA, et al. Challenges and opportunities for consistent classification of human B cell and plasma cell populations. Front Immunol 2019; 10. DOI:10.3389/fimmu.2019.02458.

26 Wrammert J, Smith K, Miller J, et al. Rapid cloning of high-affinity human monoclonal antibodies against influenza virus. Nature 2008; 453: 667–71.

27 Ellebedy AH, Jackson KJL, Kissick HT, et al. Defining antigen-specific plasmablast and memory B cell subsets in human blood after viral infection or vaccination. Nat Immunol 2016; 17: 1226–34.

28 Ramwadhdoebe TH, van Baarsen LGM, Boumans MJH, et al. Effect of rituximab treatment on T and B cell subsets in lymph node biopsies of patients with rheumatoid arthritis. Rheumatology 2019; 58: 1075–85.

29 Kamburova EG, Koenen HJPM, Borgman KJE, Ten Berge IJ, Joosten I, Hilbrands LB. A single dose of rituximab does not deplete B cells in secondary lymphoid organs but alters phenotype and function. American Journal of Transplantation 2013; 13: 1503–11.

